# Proteomic Biomarkers Differentiate Bacterial Infections in Febrile Infants: A Multicentre Prospective Study

**DOI:** 10.64898/2026.02.04.26345561

**Authors:** Clare Mills, Holly Drummond, Narainrit Karuna, Hannah Mitchell, Lisa McFetridge, Oenone Rodgers, Etimbuk Umana, Helen Groves, Thomas Waterfield

**Affiliations:** Wellcome Wolfson Institute of Experimental Medicine, Queen’s University Belfast, 97 Lisburn Road, Belfast, UK, BT9 7BL; Department of Pharmaceutical Care, Faculty of Pharmacy, Chiang Mai University, Chiang Mai, Thailand; Mathematical Sciences Research Centre, School of Mathematics & Physics, Queen’s University Belfast, Belfast, UK

**Keywords:** Febrile infants, biomarkers, proteomics, invasive bacterial infection, urinary tract infection, procalcitonin, lipocalin-2, host-response

## Abstract

**Objectives:** To identify and validate plasma host-response protein biomarkers that improve discrimination of bacterial infection in febrile infants ≤90 days of age, and to assess whether novel biomarkers add value beyond established markers.

**Methods:** Sub-study of the prospective multicentre Febrile Infant Diagnostic Assessment and Outcome (FIDO) cohort. Novel biomarkers were identified through plasma proteomic profiling (Olink®) and combined with biomarkers and signatures from the literature for verification using Luminex and ELISA platforms. Diagnostic performance of novel biomarkers, established markers (CRP, PCT), and multi-protein signatures was evaluated.

**Results:** Proteomic profiling of 110 samples identified 174 proteins differentially expressed between bacterial and viral infections, revealing distinct pathogen-specific immune signatures. Verification in the full cohort (n=445) demonstrated PCT had the highest individual accuracy for invasive bacterial infection (IBI) (AUC 0.89). Combining PCT with novel biomarkers, particularly lipocalin-2 (LCN2), improved discrimination (AUC 0.96). Diagnostic performance for the combined IBI/urinary tract infection (UTI) outcome was consistently lower (AUC <0.8).

**Conclusions:** Febrile infants demonstrate biologically coherent host-response signatures that can be leveraged diagnostically. A PCT–LCN2 combination showed excellent accuracy for identifying IBI and may support future biomarker-guided diagnostic strategies, while reliable discrimination of UTI remains challenging.

## Introduction

Febrile infants younger than 90 days of age are at high risk of urinary tract infection (UTI) and invasive bacterial infection (IBI), namely bacteraemia and meningitis (1, 2). This group presents a common diagnostic challenge, partly because they often appear well or have non-specific symptoms despite having UTI or IBI. Due to the difficulties distinguishing bacterial from non-bacterial infections in febrile infants, this cohort has traditionally been managed with a “treat-all” approach (3). While this minimises missed bacterial infections, it increases healthcare costs and exposes many infants to unnecessary invasive procedures and broad-spectrum antibiotics. Such exposure undermines antibiotic stewardship, and early-life antibiotic use may have lasting effects on health, contributing to microbiome disruption and increased risk of conditions such as obesity and asthma (4, 5).

Evidence-based clinical decision aids (CDAs) have been developed to guide the management of febrile young infants (6-9). These tools use sequential assessment based on age, clinical appearance, and host-response biomarkers to identify a subgroup of infants at low risk who may be managed without lumbar puncture or parenteral antibiotics while awaiting culture results. C-reactive protein (CRP) and procalcitonin (PCT) are the most widely used host-response biomarkers, however, their diagnostic performance is suboptimal (10-12).

Studies in older children and adults have demonstrated the value of novel protein biomarkers and multi-marker panels in distinguishing bacterial infections (13, 14). Proteins are well-suited for integration into both point-of-care diagnostics and routine laboratory workflows, with the additional advantage of requiring small sample volumes, as often obtained from infants. Despite extensive progress in infection biomarker discovery, infants are frequently underrepresented or excluded from such studies. Their distinct physiology, including immature immune responses and shorter illness duration at presentation, may critically affect biomarker performance and limit extrapolation from older populations. Additionally, blood and urine cultures, though considered the diagnostic gold standard, are often inaccurate in infants due to small sample volumes, contamination, and low pathogen yield.

This study aimed to identify and validate novel plasma protein biomarkers and multi-marker panels capable of distinguishing bacterial infection (IBI/UTI) in febrile infants ≤90 days of age, and to evaluate their diagnostic performance relative to markers currently utilised in clinical practice.

## Methods

### Study design and setting

Figure 1 shows an overview of the study procedures. This was a planned sub-study of the Febrile Infant Diagnostic Assessment and Outcome (FIDO) Study, a multicentre prospective observational cohort study that enrolled infants ≤90 days of age presenting with fever to 35 Paediatric Emergency Research in the UK and Ireland (PERUKI) sites. The study protocol, recruitment procedures, and primary results have been published previously (8, 15). The FIDO study was prospectively registered at ClinicalTrials.gov (NCT05259683; 28 February 2022). Reporting follows the Transparent Reporting of a multivariable prediction model for Individual Prognosis or Diagnosis (TRIPOD) statement (16).

**Figure 1.**
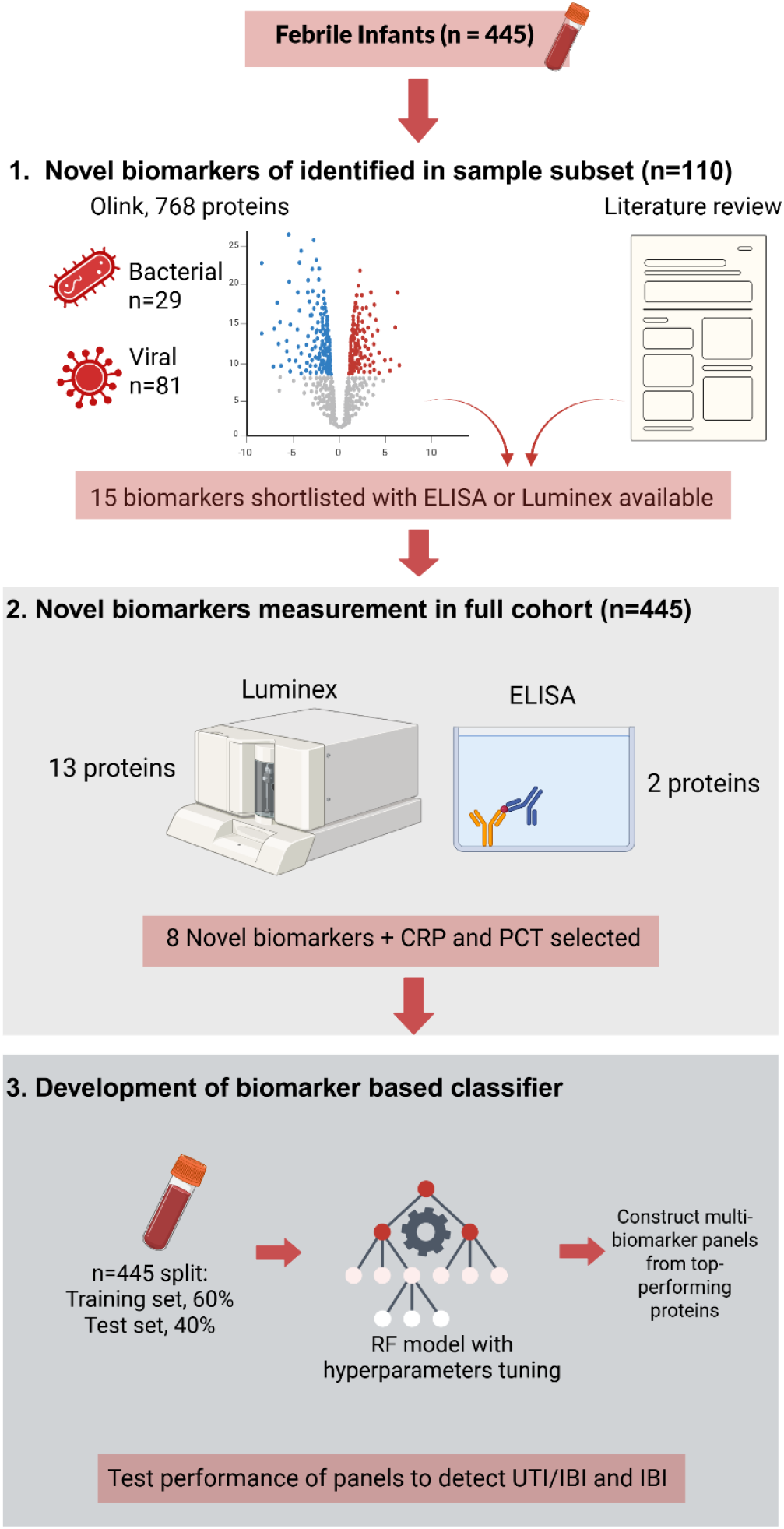
Study procedures flow chart. Figure created with BioRender.com. *CRP, C-reactive protein; IBI, Invasive bacterial infection; PCT, Procalcitonin; RF, Random forest; UTI, Urinary tract infection*.

### Participants

Infants were eligible if they were ≤90 days of age and presented with a fever ≥38 °C in the emergency department or acute unit, or if a documented fever ≥38 °C was reported within 24 h of presentation, regardless of thermometer type. Exclusion criteria were parental refusal/withdrawal of consent or missing outcome data. For this biomarker sub-study, participants were included if sufficient plasma volume was available for proteomic and biomarker analysis. Written informed consent for plasma storage and analysis was obtained from parents/guardians.

Infants were included for Olink® proteomic analysis only if there was sufficient plasma volume to permit both discovery and subsequent biomarker verification. To minimise the risk of misclassification in the discovery phase, only samples from participants with a confirmed viral pathogen were included for comparison with bacterial infections; samples in which no pathogen was identified were excluded.

### Data Collection and Definitions

Case report forms (CRFs) were prospectively completed at first clinical assessment, prior to the availability of laboratory results. “Unwell appearance” was defined as abnormal global assessment and/or abnormal age-specific vital signs (as per APLS recommendations). A follow-up CRF was completed 7 days after discharge to capture investigation results and clinical outcomes.

### Outcome

The outcome of bacterial infection was defined as UTI and IBI combined. All outcomes were determined using routine microbiological testing conducted at accredited NHS laboratories, independently of the study investigators. All testing was conducted on samples collected at initial presentation. Cut-off values and lists of pathogens/contaminants were predefined (15).

Invasive bacterial infection (IBI): Defined as culture or molecular detection of a bacterial pathogen in blood or cerebrospinal fluid (CSF).

Urinary tract infection (UTI): Defined by any of the following:

- Growth of a single organism ≥10^5^ CFU/mL in clean-catch urine.
- Growth of a single organism ≥10^4^ CFU/mL from transurethral bladder catheter (TUBC) or suprapubic aspiration (SPA).
- Growth of the same organism ≥10^5^ CFU/mL from ≥2 pad/bag samples plus pyuria.

Pyuria: Defined as leucocyte esterase ≥trace on dipstick or ≥10 WBC/hpf on centrifuged urine microscopy.

Viral infection: Defined as the detection of a virus in the CSF or on a respiratory viral test. This included both laboratory-based and POC tests.

### Olink® proteomic profiling and data analysis

Plasma protein levels (n = 768) were quantified using Olink® Explore panels (“Neurology” and “Inflammation”) at an Olink® Certified Service Provider (Randox, UK). Data were normalised and log2-transformed (Normalised Protein Expression; NPX). Proteins detected in <50% of samples or duplicated across panels were excluded, leaving 724 proteins.

Olink® data were analysed using R (v4.3.3). Proteins differentially expressed between infection groups were identified with the “Olink® Analyze” R package. Statistical significance was determined using a false discovery rate (FDR) threshold of <0.05, corrected by the Benjamini– Hochberg method. For heatmap visualisation, protein values were first scaled using the scale() function. Hierarchically clustered heatmaps were then generated with the ComplexHeatmap package, incorporating clinical annotations via HeatmapAnnotation().

Functional enrichment of differentially expressed proteins was performed using the Metascape Gene Annotation and Analysis Resource (http://metascape.org), querying KEGG, Gene Ontology (GO), and Reactome gene sets (17). Enrichment analysis used the following criteria: minimum count ≥3, enrichment factor >1.5, and p <0.01. The full list of proteins measured on the Olink® panels was included as a background list. Significantly enriched terms were automatically grouped into non-redundant clusters by Metascape and presented as bar charts.

### Predictors for biomarker models

Clinical and routine biomarkers: CRP testing was conducted at local accredited NHS laboratories as part of the child’s routine investigations on samples collected at initial presentation, as part of routine care by staff blind to clinical assessment data. PCT was measured at the Department of Clinical Biochemistry, Royal Victoria Hospital Belfast, using the Elecsys BRAHMS assay (Thermo Fisher) on the Roche e801. Laboratory staff were blinded to clinical and outcome data.

Additional biomarkers: Biomarkers from Olink® analysis were selected as predictors for evaluation in the full biomarker cohort if they met the criteria of adjusted P-value <0.01 and fold change >0.5 between bacterial and viral groups, and if they were available on multiplex platforms suitable for the small sample volumes obtainable from infants. Additional biomarkers were identified from a recent systematic review of bacterial versus viral biomarkers in older children and adults (18).

The additional biomarkers were measured on Luminex and ELISA immunoassays were performed at Queen’s University Belfast by laboratory staff blinded to clinical and outcome data. Luminex was used to quantify C-C motif chemokine ligand 20 (CCL20), C-C motif chemokine ligand 23 (CCL23), E-Selectin, interferon-gamma (IFN-γ), interleukin-6 (IL-6), interleukin-17A (IL-17A), interleukin-18 (IL-18), interferon gamma-induced protein 10 (IP-10), matrix metalloproteinase-8 (MMP8), neural cell adhesion molecule 1 (NCAM1), TNF-related apoptosis-inducing ligand (TRAIL), TNF-related apoptosis-inducing ligand receptor 2 (TRAILR2), and TNF-related activation-induced cytokine (TRANCE) using the Luminex® Discovery Assay (Bio-Techne, Abingdon, UK) on a Luminex® 100/200™ analyser, following the manufacturer’s protocol. Batch effects were corrected using ComBat (19). LCN2 and LG3BP were measured on commercial ELISA kits (Quantikine® ELISA kit, Bio-techne, Abingdon, UK), as per manufacturer’s protocol. For both ELISA and Luminex assays, values that were out of range were replaced by the closest calibrator curve point and samples were randomised across plates.

### Sample size

Sample size estimation was performed to ensure adequate precision in the estimated discriminative ability of the model for bacterial infection (UTI/IBI), expressed as receiver operating characteristic (ROC) curves, with area under the curve (AUC) values. Calculations were conducted using the powerROC web tool (18). Inputs included an anticipated AUC of 0.90, a disease prevalence of 15%, and a target 95% confidence interval width of less than 0.10 (i.e., precision of ±0.05). Under these assumptions, the minimum required sample size was 308 participants, including approximately 46 events. The final study population comprised 445 infants, which exceeds this requirement and provides sufficient precision for estimation of the model’s discriminative performance for bacterial infection.

To account for the limited number of IBI events and mitigate the risk of overfitting, we restricted the predictor set and applied penalisation within a cross-validation framework. Class imbalance was further addressed using the Synthetic Minority Oversampling Technique (SMOTE), consistent with TRIPOD methodological recommendations.

### Biomarker models

Biomarkers were included as predictors for random forest (RF) classifier models if they were statistically different between IBI/UTI and no bacterial infection. RF classifier was implemented in R program (v4.4.3) (*tidymodels* version 1.2; https://CRAN.R-project.org/package=tidymodels). Data were split into training (60%) and test (40%) sets with stratification by IBI or IBI/UTI status. Model development was conducted on the training set using five-fold cross-validation, with class imbalance addressed by SMOTE algorithm (over_ratio = 0.5). Hyperparameters (*mtry, min_n*) were optimised using grid search, and model performance was assessed by ROC AUC. The final model was refit with the optimal parameters and evaluated on the independent test set. Proteins demonstrating high discriminatory performance (AUC > 0.80) were systematically combined with an established clinical biomarker to construct candidate panels with improved diagnostic performance. In parallel, feature selection using FS-PLS was also explored, followed by multivariable binary logistic regression modelling.

### Statistical analysis

All statistical assumptions were evaluated before conducting the analysis. Categorical clinical variables were summarised as frequencies and percentages, with group differences assessed using Fisher’s exact test. Continuous variables were reported as mean ± standard deviation (SD) for normally distributed data, as determined by the Shapiro–Wilk test, or as median and interquartile range (IQR) for non-normally distributed data. Group comparisons were performed using Student’s t-test for normally distributed variables or the Mann–Whitney U-test for non-normally distributed variables. Statistical significance was defined as *p* <0.05. Participants with missing outcome or biomarker data were excluded from relevant analyses.

The diagnostic accuracy of single biomarkers and previously published signatures was evaluated using ROC curves, with AUC values and 95% confidence intervals calculated using R program (v4.3.3) with the pROC package. For previously published signatures, disease risk scores (DRS) were generated as described below, as per the original studies (20, 21). The Jackson et al weighted DRS (E-selectin+IL18+NCAM1+LCN2+IFN-γ) and Jackson et al simple DRS (E-selectin+IL18+NCAM1+LCN2+IFN-γ+LG3BP) signatures were strictly reproduced, while the Oved et al signature (TRAIL+IP-10+CRP) was recalibrated to this cohort to account for differences in biomarker measurement platforms. For the Jackson et al weighted DRS, protein expression values were log2-transformed prior to calculation and values for each NCAM1, IFN-γ, E-Selectin, LCN2, and IL-18 were then multiplied by the published coefficients, with the weighted sum used as the DRS value. For the Jackson et al simple DRS, protein expression values were log2-transformed and then calculated as (LCN2 + E-Selectin + IFN-γ) – (LG3BP + IL-18 + NCAM1). The Oved et al signature was assessed using multivariable binary logistic models. Stepwise selection was first used to determine the optimal model for both bacterial infection (UTI/IB) and IBI alone.

### Study ethics

Ethical approval was obtained from the Office for Research Ethics Committees Northern Ireland (22/NI/0002), the Public Benefit and Privacy Panel for Health and Social Care Scotland (2122-0257), and the Children’s Health Ireland Research and Ethics Committee (REC-082-22).

## Results

### Patient Characteristics

Plasma samples for biomarker testing were available for 445 FIDO study participants, of whom 64/445 (14.3%) had a bacterial infection (UTI/IBI) and 23/445 (5.2%) had an IBI alone. The flow of participants through the study is shown in Supplemental Figure 1, and Table 1 compares the baseline characteristics of the total FIDO cohort from the parent study with those included in this sub-study (biomarker cohort). There were no significant differences in baseline characteristics between the two groups, except that infants in the biomarker cohort were more frequently reported as having an unwell appearance (292/445 [65.6%]) compared with the total cohort (1058/1821 [58.1%]). Consistent with this observation, infants in this biomarker cohort were more likely to receive antibiotics, be admitted to hospital, and receive a diagnosis of bacterial infection (UTI/IBI). The rate of bacterial infection (UTI/IBI) was slightly but significantly higher in the biomarker cohort than in the total cohort (14.4% vs 10.8%, *p* = 0.04). IBI rates were also higher in the biomarker cohort, although this difference did not reach statistical significance (5.2% vs 3.7%, *p* = 0.15). The distribution of bacterial pathogen species was similar between the total and sub-study biomarker cohorts (Supplemental Table 1).

**Table 1.** Demographic and clinical characteristics of full FIDO cohort and those available for additional biomarker testing. *IQR; Interquartile range, IBI; Invasive Bacterial Infection, UTI: Urinary Tract Infection, LOS; Length of Stay*.

| Variable | Biomarker cohort | Total study cohort | P-value |
| --- | --- | --- | --- |
| **N** | 445 | 1821 |  |
| **Baseline characteristics** |  |  |  |
| **Age (days), Median (IQR)** | 47 (30 -66) | 46 (30-64) | 0.495 |
| **Male, n(%)** | 276 (62.0) | 1108 (60.8) | 0.687 |
| **Comorbidities present, n(%)** | 65 (14.6) | 260 (14.3) | 0.859 |
| **Presenting <6 hours after fever onset, n(%)** | 239 (53.7) | 966 (53.0) | 0.802 |
| **Unwell appearance, n(%)** | 292 (65.6) | 1058 (58.1) | 0.004 |
| **Temperature (°C), Median (IQR)** | 38 (37.4-38.4) | 38 (37.4-38.4) | 0.560 |
| **Outcomes** |  |  |  |
| **Antibiotics prescribed, n(%)** | 391 (87.9) | 1242(68.2) | <0.001 |
| **Admitted to ward, n(%)** | 363 (81.6) | 1407 (77.2) | 0.048 |
| **LOS (days), n(%)** | 2 (2-4) | 2 (2-3) | 0.248 |
| **UTI/IBI, n(%)** | 64 (14.4) | 198 (10.8) | 0.037 |
| **IBI, n(%)** | 23 (5.2) | 67 (3.7) | 0.149 |

### Proteomic biomarker profiling

Of the 445 available samples, 110 (29 bacterial, 81 viral) were selected for Olink® discovery (Figure 1, Supplemental Figure 1). Characteristics and infection details of those included in the Olink® cohort are shown in Supplemental Table 2. A total of 174 proteins were differentially expressed between bacterial and viral groups (FDR < 0.05) (Figure 2A, Supplemental Table 3). Enrichment analysis of proteins significantly (FDR<0.05) higher in bacterial or viral infections highlighted clear aetiology-specific biology (Figure 2B). In infants with bacterial infection, enriched pathways included response to bacterium, regulation of macrophage activation, and regulation of myeloid cell differentiation, consistent with a strong innate immunity-driven response. There was also clear upregulation of classical neutrophil granule enzymes (MPO, MMP8, FCAR) in bacterial cases (Figure 2A, Supplemental Table 3). In viral infection, enriched categories included “defence response to virus,” driven by the proteins IFNL1, IFNLR1, BST2, TRIM21, TLR3, CXCL10, and CXCL11, consistent with activation of interferon-mediated antiviral defences.

**Figure 2.**
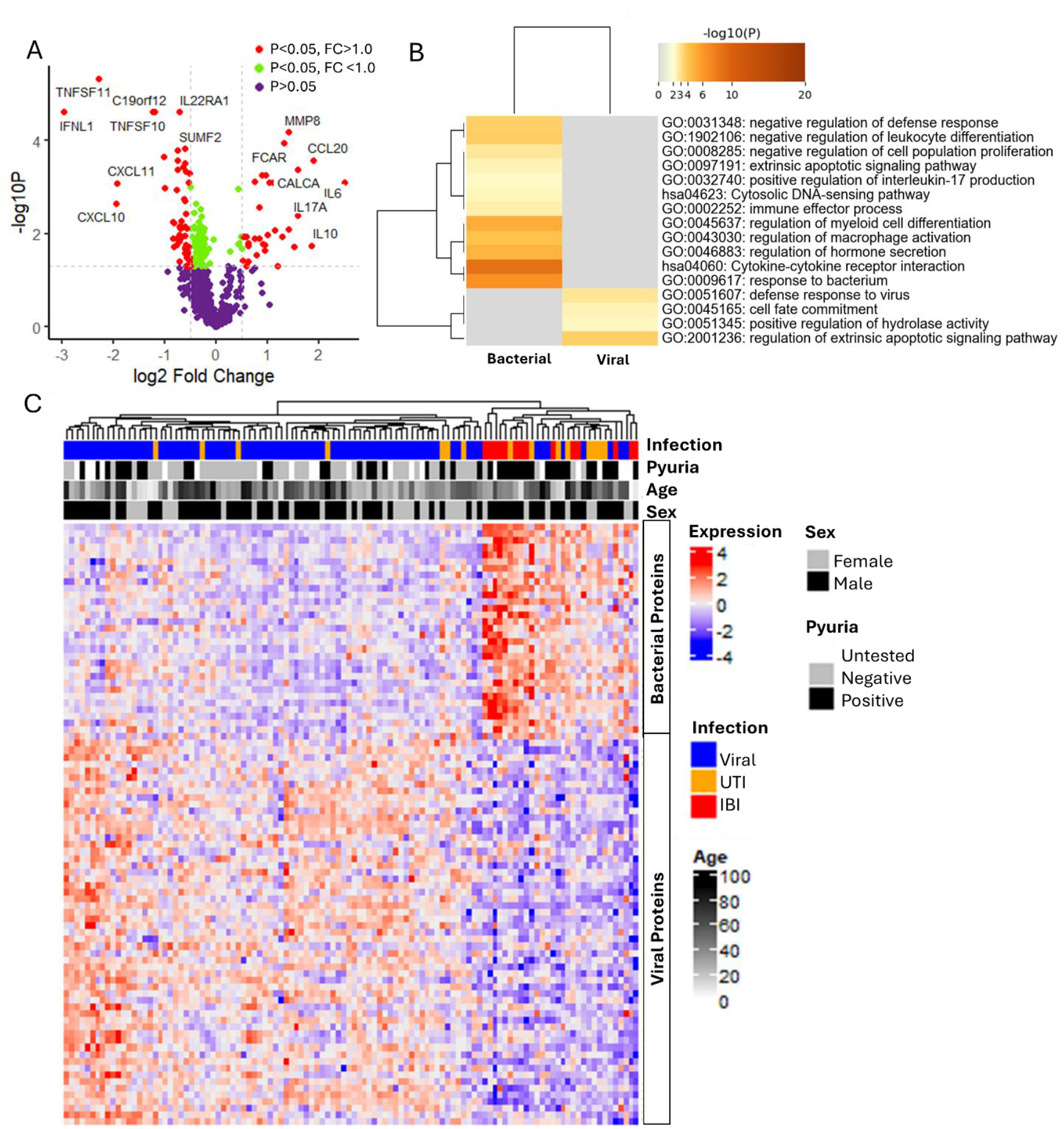
Host plasma proteome response to bacterial and viral infections in infants. (A) Volcano plot of differentially expressed proteins (FDR <0.05) between infants with bacterial infection (UTI/IBI, n=29) and viral infection (n=81) following Olink® proteomic profiling. (B) Bar chart of enriched terms from proteins significantly (FDR <0.05) up and down regulated in infants with bacterial or viral infections. Coloured by p-values, metascape. (C) Hierarchical clustering displayed as a heatmap of significantly differentiated proteins (FDR<0.05) with a fold change of >0.5 between bacterial and viral samples measured across the Olink cohort (columns). Scaled expression values are shown (red = higher, blue = lower relative abundance). Annotation bars indicate infection (IBI, UTI or viral), age in days, sex, or presence of pyuria. *IBI; Invasive bacterial infection; FC, Fold change; FDR, False discovery rate; UTI; Urinary tract infection*,

A hierarchically clustered heatmap of differentially expressed proteins (FDR<0.05, FC >0.5) is shown in Figure 2C. Clustering revealed a clear distinction between bacterial (IBI/UTI) and viral infections. Most IBI cases were grouped within a distinct bacterial cluster, while a small number of viral cases clustered alongside these, displaying inflammatory proteomic profiles more typical of bacterial infection. UTI cases showed greater heterogeneity, with some aligning with IBI and others with viral infections. UTI samples without pyuria tended to cluster with viral infections and demonstrated up-regulation of viral-associated proteins, whereas those with pyuria aligned more closely with the bacterial cluster.

### Diagnostic accuracy of novel biomarkers

Proteins from the Olink® discovery analysis showing the strongest differential expression (FDR < 0.01, FC > 0.5) and suitability for multiplex quantification were prioritised for verification in the total biomarker cohort. Selection also considered platform compatibility with small infant plasma volumes, and additional candidate biomarkers were incorporated based on evidence from a recent systematic review of bacterial versus viral infection biomarkers in older paediatric and adult populations (18).

The selected proteins were measured in the full cohort (n=445) using Luminex and ELISA platforms, which are more clinically applicable than Olink®. In total, 15 proteins were assayed: 8 were elevated in bacterial infection and 7 were decreased (Supplemental Table 4). IL-17 was subsequently excluded as values were below the limit of detection in 98.4% of samples. Box-and-whisker plots for all proteins comparing UTI/IBI vs. no bacterial infection and IBI vs. no IBI are shown in Supplemental Figures 2 and 3, respectively.

**Figure 3.**
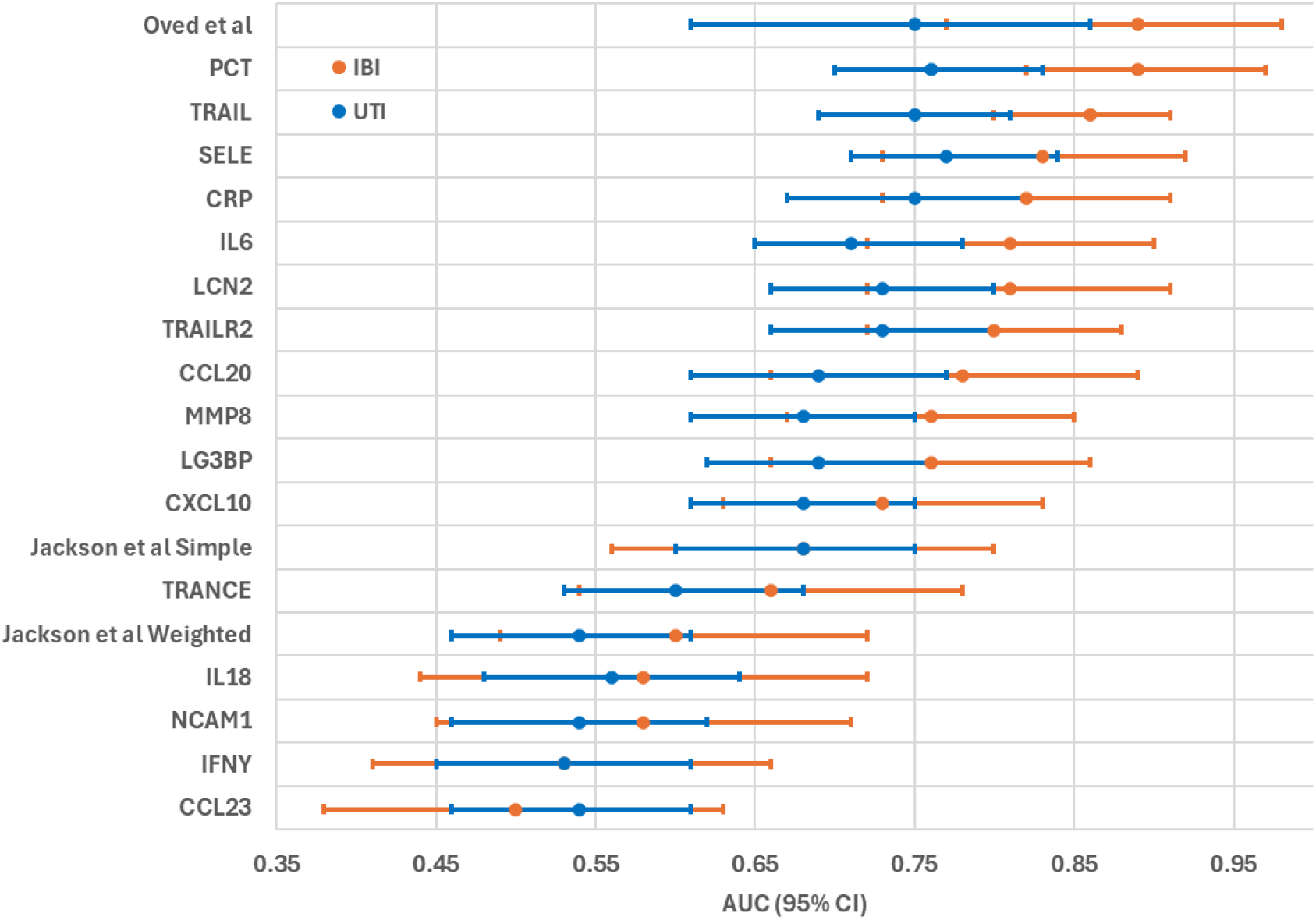
Diagnostic accuracy of biomarkers for IBI and IBI/UTI. AUC values with 95% confidence intervals for each individual biomarker or previously published signature. Blue markers indicate IBI vs no IBI; orange markers indicate IBI/UTI vs no bacterial infection. *AUC; Area Under the Curve, UTI; Urinary tract infection, IBI; Invasive bacterial infection*.

Supplemental Table 5 and Figure 3 summarise the diagnostic performance of the candidate biomarkers alongside three previously published biomarker signatures (Jackson et al weighted; Jackson et al simple; and Oved et al.). The Oved et al. signature was modelled according to the original publication (logistic regression) but was recalibrated to this cohort to account for differences in biomarker measurement platforms. Based on stepwise selection, the optimal model included only TRAIL and CRP for both the IBI and UTI/IBI comparisons. The model outputs are presented in Supplemental Table 6.

For discriminating IBI/UTI from no bacterial infection, PCT (AUC 0.76, 95% CI 0.70–0.83), the Oved et al signature (AUC 0.78, 95% CI 0.72–0.84), and SELE (AUC 0.77, 95% CI 0.71–0.84) achieved the highest accuracy. No single biomarker or published multi-marker signature achieved high discrimination (AUC>0.8) for IBI/UTI. Biomarker AUC values for distinguishing IBI from no IBI were considerably higher. For IBI vs no IBI several biomarkers outperformed CRP, but none exceeded PCT (AUC 0.89, 95% CI 0.82–0.97). The Oved et al signature performed comparably (AUC 0.89, 95% CI 0.83–0.95), followed closely by TRAIL (AUC 0.86, 95% CI 0.80– 0.91).

### Development of an optimal biomarker signature

We next developed multivariable diagnostic models to assess whether combining established and novel biomarkers improved discrimination of bacterial infection. A RF model was used to evaluate the discriminatory performance of individual biomarkers in distinguishing UTI/IBI or IBI from controls. The 12 proteins that were significantly different between IBI/UTI and no bacterial infection (Supplemental Figure 2) were assessed individually, with discrimination quantified by fivefold cross-validation and expressed as AUC (Figure 4a). For discriminating IBI, consistent with univariate ROC analyses, PCT demonstrated the highest individual performance (AUC=0.90).

**Figure 4.**
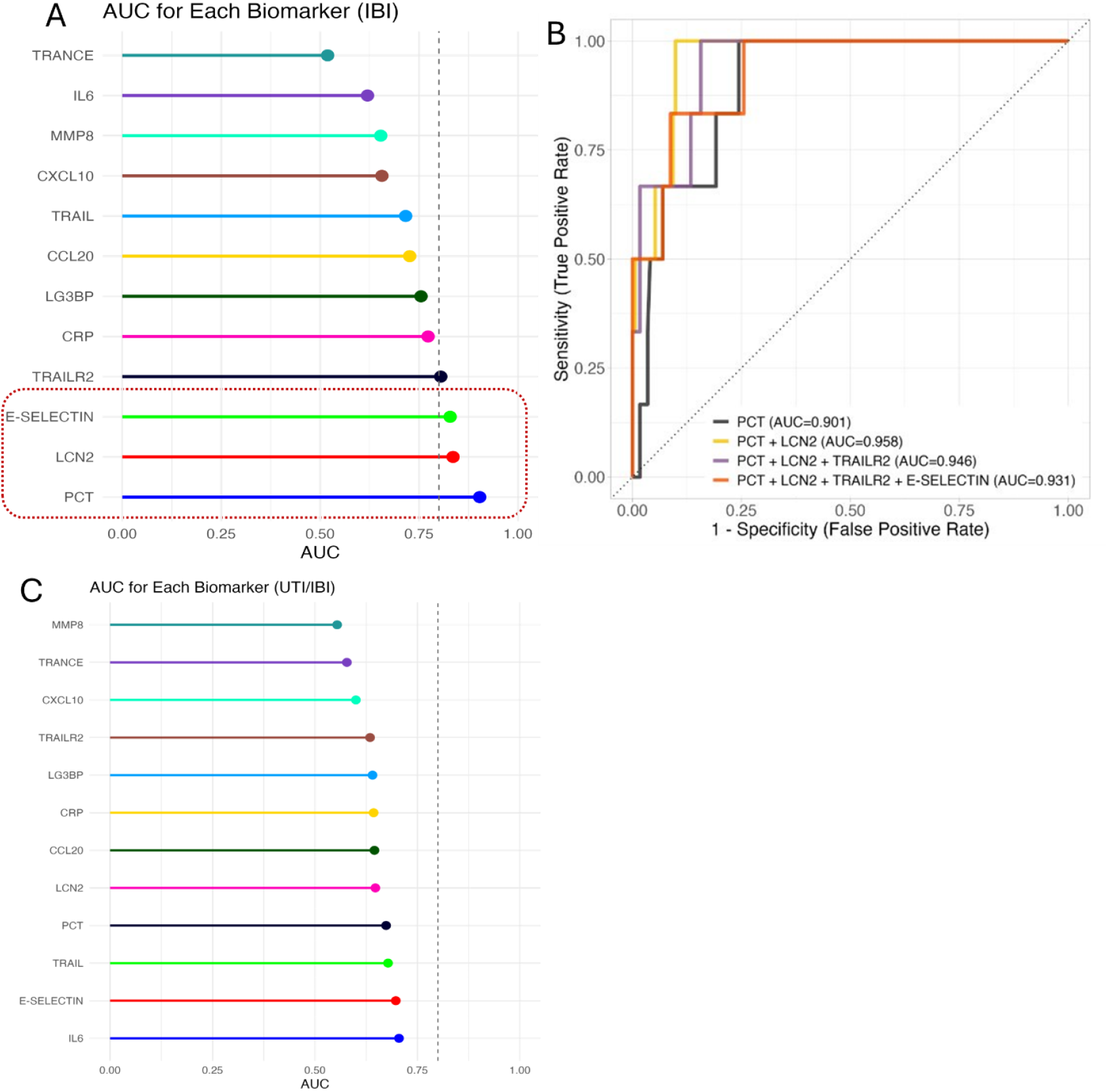
Diagnostic performance of multi-biomarker panels. (A) AUC values each individual biomarkers for IBI following RF model. (B) ROC curve showing comparison of AUC values for PCT alone versus PCT combined with top-performing novel biomarkers for IBI. (C) AUC values each individual biomarkers for UTI following RF model. *AUC, Area Under the Curve; IBI, Invasive bacterial infection; PCT, Procalcitonin; RF, Random Forest; ROC, Receiver Operating Characteristic; UTI, Urinary tract infection*

To determine whether novel proteins added value beyond established markers, multi-biomarker panels were constructed by combining PCT with the top-performing proteins (AUC >0.80) (Figure 4b). All combinations improved discrimination compared with PCT alone, with multi-biomarker panels achieving AUCs >0.90. The highest diagnostic performance was observed for the PCT+LCN2 combination (AUC=0.96). The same approach was applied to the discrimination of UTI/IBI; however, no biomarker combination achieved an AUC of >0.80 (Figure 4C).

Given the more limited performance of RF models for UTI/IBI, we further investigated alternative feature selection and modelling approaches. We applied forward selection–partial least squares (FS-PLS) followed by multivariable logistic regression, which identified CCL20, IP-10, TRAIL, IL-6, and CCL23 as candidate predictors. Stepwise selection subsequently removed IL-6. The resulting model achieved an AUC (95% CI) of 0.79 (0.66–0.90), indicating modest discrimination that only moderately exceeded the RF-derived combinations.

## Discussion

In this multicentre prospective study of febrile infants ≤90 days of age, plasma proteomic profiling was used to identify and validate novel biomarkers that distinguish bacterial from viral infections with high diagnostic accuracy. Biomarker AUC values were considerably higher for distinguishing IBI rather than IBI/UTI from non-bacterial infection. Established marker PCT demonstrated the highest diagnostic performance among individual biomarkers for distinguishing IBI in febrile infants (AUC = 0.89), while the Oved et al signature (TRAIL+IP-10+CRP) exhibited comparable discrimination (AUC = 0.89). Notably, combining PCT with novel host-response proteins, particularly LCN2, further improved diagnostic accuracy to distinguish IBI (AUC = 0.96), highlighting the potential clinical utility of this multi-marker approach for early risk stratification in this vulnerable population.

The performance of the PCT + LCN2 panel may reflect the incorporation of PCT, an acute phase reactant, and LCN2, a rapid indicator of neutrophil activation and bacterial iron sequestration (22). LCN2’s biological properties make it particularly suitable as an early infection marker in infants, who often present shortly after symptom onset. Unlike CRP and PCT, which require de novo synthesis, LCN2 is pre-formed and is released within an hour of neutrophil activation in stimulated blood samples, suggesting it may rise earlier in infection, though its in vivo plasma kinetics remain to be defined (23).

Previously reported diagnostic accuracies (AUCs) for LCN2 in discriminating bacterial infection vary widely, from 0.5 to 0.97, reflecting heterogeneity in study populations, infection definitions, and assay platforms (20-22). Structurally, LCN2 exists in two major forms: a monomeric 22 kDa protein, produced by epithelial cells under inflammatory conditions, and a dimeric 45 kDa protein, predominantly neutrophil-derived (23). In the present study, total LCN2 (both forms) were quantified. However, assays that specifically measure the dimeric form have demonstrated improved specificity for bacterial infection, with AUC increasing from 0.89 to 0.97 (20). Future work should assess dimer-specific LCN2 measurement in infants, which may further enhance diagnostic accuracy.

Importantly, we evaluated the diagnostic performance of previously reported high-performing host-response signatures in infants. The Oved et al. signature (TRAIL + CRP + IP-10) performed well for IBI in our cohort (AUC = 0.89), outperforming CRP alone (AUC = 0.82) but remaining comparable to PCT (AUC = 0.89). Consistent with our findings, the Oved et al. signature has shown high diagnostic accuracy across multiple studies (24-27). Notably, this signature is available commercially as the MeMed assay. However, due to sample volume constraints, TRAIL, CRP, and IP-10 were measured on an alternative platform rather than using the proprietary MeMed assay. Recognising this and that biomarker cut-offs and regression coefficients likely differ in infants, we recalibrated the logistic regression model using our dataset. Interestingly, IP-10 did not contribute significantly to model performance. While the Oved et al. signature showed high diagnostic performance, it did not surpass PCT in our cohort, and further optimisation, potentially excluding IP-10, may be required for application in young infants. In contrast, both the Jackson et al weighted and simple DRS signatures performed poorly in our cohort, indicating limited generalisability to this age group.

Comprehensive proteomic analysis identified 174 proteins differentially expressed between bacterial and viral infections, revealing clear pathogen-specific immune responses even in very young infants. Bacterial infections were characterised by strong activation of innate immune pathways, consistent with neutrophil-driven inflammation observed in older children and adults (28). Conversely, viral infections demonstrated enrichment of established interferon-mediated antiviral defence pathways (29). These findings confirm that, despite immune immaturity, febrile infants exhibit distinct and biologically coherent host-response signatures that can be leveraged for diagnostic purposes, as reflected by the improved diagnostic performance achieved with novel biomarkers for IBI.

Conversely, all individual biomarkers and multi-marker signatures performed poorly for the combined UTI/IBI outcome, with no model achieving an AUC greater than 0.8. Hierarchical clustering of proteins identified from the proteomic analysis showed that IBI cases clustered tightly within the bacterial expression profile, while UTI samples displayed greater heterogeneity, some aligning with bacterial, others with viral responses. We therefore hypothesise that this reduced diagnostic accuracy primarily reflects misclassification of UTI cases rather than limitations in biomarker performance due to an attenuated systemic response to UTI. Notably, UTI samples lacking pyuria tended to cluster with upregulation of viral responses, whereas those with pyuria grouped more consistently within the bacterial cluster. There remains considerable debate regarding whether pyuria should be a required criterion for defining UTI in infants (30). This controversy reflects the high risk of contamination inherent to infant urine collection methods and evidence that some bacterial pathogens, particularly Gram-positive species, elicit a weaker pyuric response than *Escherichia coli* (31). The definition of UTI from the parent FIDO study did not include pyuria, and we retained this approach to avoid incorporation bias when evaluating systemic host-response biomarkers (15). However, our proteomic data suggest that this definition may inadvertently include infants with viral infections and contaminated urine cultures. Although the number of UTI cases precluded a definitive statistical assessment, these findings highlight the need for further work integrating host-response profiling, pyuria status, and pathogen detection to refine UTI diagnostic definitions in infants.

This study has several important strengths. It represents one of the largest multicentre, prospectively recruited cohorts of febrile infants to undergo systematic biomarker evaluation. Rigorous case definitions, blinded laboratory analyses, and a predefined analytic protocol enhance internal validity. The inclusion of PCT, rather than CRP alone, represents a more meaningful benchmark comparator. Given PCT’s consistently higher diagnostic accuracy but limited clinical uptake due to cost and uncertain impact on outcomes, its use provides a more stringent and meaningful standard for comparison (11, 32). This approach underscores the potential clinical relevance of biomarkers that can outperform PCT, not merely CRP.

Despite the limited plasma volumes obtainable from infants, we successfully integrated both proteomic discovery and targeted validation phases, incorporating established and novel biomarkers identified from the literature. The use of clinically relevant multiplex and ELISA platforms improves translational applicability. Model development adhered to TRIPOD recommendations, including internal cross-validation and calibration assessment, to minimise overfitting.

Several limitations should also be acknowledged. Although baseline characteristics suggest that the biomarker cohort was broadly representative of the full FIDO population, infants included in this sub-study were more often unwell-appearing and had higher rates of bacterial infection, likely reflecting sampling bias toward those from whom sufficient plasma could be obtained. This may partly explain the high discriminative performance of PCT observed in our study, compared to others, as more severely ill infants tend to have stronger systemic responses (11). Consequently, the generalisability of our findings to well-appearing infants remains to be confirmed through external validation.

Finally, while the study was adequately powered for bacterial infection overall (IBI/UTI), diagnostic improvement was primarily observed for IBI. This likely reflects limitations in the reference standard for UTI diagnosis in infants rather than a true lack of biomarker performance. IBI, however, represents the most clinically consequential infection category in this population, reinforcing the relevance of our findings to current diagnostic and management challenges. The number of IBI cases was modest, reflecting the low incidence of such infections in this age group and limiting model complexity and precision of estimates. Nonetheless, the study screened over 2,000 infants across 35 sites to achieve the final cohort size, underscoring the feasibility and scalability of this approach for future validation studies.

In summary, this multicentre prospective study identified and validated plasma protein biomarkers that distinguish IBI in febrile infants with high diagnostic accuracy. The combination of PCT and LCN2 demonstrated excellent diagnostic accuracy within our RF model, suggesting potential utility pending further clinical validation. These findings provide new insights into the host immune response in early infancy and highlight the potential for biomarker-guided diagnostics to improve care and antibiotic stewardship. Future work should focus on external validation, evaluation in well-appearing infants, and integration of biomarker-based approaches into clinical decision aids and guideline frameworks to support safer, evidence-based management of febrile infants.

## Acknowledgements

We thank the funders of this study, Royal College of Emergency Medicine Doctoral Fellowship (RCEM 02/03/2021) and Sir Halley Stewart Trust (4292). We also thank the children and their families who participated in the FIDO study, and the staff at the FIDO study hospital sites who participated in screening and enrolment

## Author contributions

Dr Clare Mills, Dr Etimbuk Umana, Dr Helen Groves, and Dr Tom Waterfield conceived and designed the study. Dr Tom Waterfield served as Principal Investigator of the FIDO study, secured study funding, and provided overall supervision and critical manuscript review. Dr Etimbuk Umana coordinated and led the running of the original FIDO study. Dr Clare Mills oversaw Olink® sample processing and proteomic data analysis, supervised ELISA and Luminex biomarker assays, verified individual biomarker analyses, and drafted the initial manuscript. Dr Holly Drummond performed ELISA and Luminex biomarker assays, contributed to individual biomarker data analysis. Dr Narainrit Karuna designed and implemented the machine-learning modelling, including random forest (RF) biomarker modelling, and contributed to the interpretation of the modelling outputs. Dr Hannah Mitchell and Dr Lisa McFetridge conducted FS-PLS and multivariable logistic regression analyses. Oenone Rodgers performed PCT assays and contributed to plasma sample collection. All authors contributed to data interpretation, reviewed, and approved the final manuscript.

## Data availability

Data available upon reasonable request.

## AI Use Statement

Artificial intelligence (ChatGPT by OpenAI) was used to assist with improving the clarity, grammar, and readability of the manuscript. All AI-generated content was subsequently reviewed and edited by all authors to ensure scientific accuracy and integrity. No AI tools were used for data analysis or interpretation.

## References

1. Ladhani SN, Henderson KL, Muller-Pebody B, Ramsay ME, Riordan A. Risk of invasive bacterial infections by week of age in infants: prospective national surveillance, England, 2010-2017. Arch Dis Child. 2019;104(9):874–8.

2. Hunt KM, Green RS, Sartori LF, Aronson PL, Chamberlain JM, Florin TA, et al. Urine Dipstick for the Diagnosis of Urinary Tract Infection in Febrile Infants Aged 2 to 6 Months. Pediatrics. 2025;155(4).

3. Craig JC, Williams GJ, Jones M, Codarini M, Macaskill P, Hayen A, et al. The accuracy of clinical symptoms and signs for the diagnosis of serious bacterial infection in young febrile children: prospective cohort study of 15 781 febrile illnesses. BMJ. 2010;340:c1594.

4. Reyman M, van Houten MA, Watson RL, Chu MLJN, Arp K, de Waal WJ, et al. Effects of early-life antibiotics on the developing infant gut microbiome and resistome: a randomized trial. Nature Communications. 2022;13(1):893.

5. Duong QA, Pittet LF, Curtis N, Zimmermann P. Antibiotic exposure and adverse long-term health outcomes in children: A systematic review and meta-analysis. Journal of Infection. 2022;85(3):213–300.

6. Gomez B, Mintegi S, Bressan S, Da Dalt L, Gervaix A, Lacroix L. Validation of the “Step-by-Step” Approach in the Management of Young Febrile Infants. Pediatrics. 2016;138(2).

7. Waterfield T, Lyttle MD, Munday C, Foster S, McNulty M, Platt R, et al. Validating clinical practice guidelines for the management of febrile infants presenting to the emergency department in the UK and Ireland. Arch Dis Child. 2022;107(4):329–34.

8. Umana E, Mills C, Norman–Bruce H, Mitchell H, McFetridge L, Lynn F, et al. Performance of clinical decision aids (CDA) for the care of young febrile infants: a multicentre prospective cohort study conducted in the UK and Ireland. eClinicalMedicine. 2024;78.

9. Pantell RH, Roberts KB, Adams WG, Dreyer BP, Kuppermann N, O’Leary ST, et al. Evaluation and Management of Well-Appearing Febrile Infants 8 to 60 Days Old. Pediatrics. 2021;148(2).

10. Borowski S, Shchors I, Bar-Meir M. Time from symptom onset may influence C-reactive protein utility in the diagnosis of bacterial infections in the NICU. BMC Pediatrics. 2022;22(1):715.

11. Norman-Bruce H, Umana E, Mills C, Mitchell H, McFetridge L, McCleary D, et al. Diagnostic test accuracy of procalcitonin and C-reactive protein for predicting invasive and serious bacterial infections in young febrile infants: a systematic review and meta-analysis. Lancet Child Adolesc Health. 2024;8(5):358–68.

12. Abe R, Oda S, Sadahiro T, Nakamura M, Hirayama Y, Tateishi Y, et al. Gram-negative bacteremia induces greater magnitude of inflammatory response than Gram-positive bacteremia. Crit Care. 2010;14(2):R27.

13. Leticia Fernandez-Carballo B, Escadafal C, MacLean E, Kapasi AJ, Dittrich S. Distinguishing bacterial versus non-bacterial causes of febrile illness - A systematic review of host biomarkers. J Infect. 2021;82(4):1–10.

14. Gunaratnam LC, Robinson JL, Hawkes MT. Systematic Review and Meta-Analysis of Diagnostic Biomarkers for Pediatric Pneumonia. J Pediatric Infect Dis Soc. 2021;10(9):891–900.

15. Umana E, Mills C, Norman-Bruce H, Wilson K, Mitchell H, McFetridge L, et al. Applying clinical decision aids for the assessment and management of febrile infants presenting to emergency care in the UK and Ireland: Febrile Infant Diagnostic Assessment and Outcome (FIDO) Study protocol. BMJ Open. 2023;13(9):e075823.

16. Moons KG, Altman DG, Reitsma JB, Ioannidis JP, Macaskill P, Steyerberg EW, et al. Transparent Reporting of a multivariable prediction model for Individual Prognosis or Diagnosis (TRIPOD): explanation and elaboration. Ann Intern Med. 2015;162(1):W1–73.

17. Zhou Y, Zhou B, Pache L, Chang M, Khodabakhshi AH, Tanaseichuk O, et al. Metascape provides a biologist-oriented resource for the analysis of systems-level datasets. Nature Communications. 2019;10(1):1523.

18. Drummond H, Mills C, Groves H, Waterfield T. Novel biomarkers for distinguishing bacterial from non-bacterial infection: a systematic review. medRxiv. 2025:2025.09.03.25334997.

19. Leek JT, Johnson WE, Parker HS, Jaffe AE, Storey JD. The sva package for removing batch effects and other unwanted variation in high-throughput experiments. Bioinformatics. 2012;28(6):882–3.

20. Yu Z, Jing H, Hongtao P, Furong J, Yuting J, Xu S, et al. Distinction between bacterial and viral infections by serum measurement of human neutrophil lipocalin (HNL) and the impact of antibody selection. J Immunol Methods. 2016;432:82–6.

21. Wang Y, Zhang M, Huang M, Wang T, Wei W, Yin B, et al. Detection of serum human neutrophil lipocalin is an effective biomarker for the diagnosis and monitoring of children with bacterial infection. Diagn Microbiol Infect Dis. 2023;106(2):115943.

22. Esposito S, Bianchini S, Gambino M, Madini B, Di Pietro G, Umbrello G, et al. Measurement of lipocalin-2 and syndecan-4 levels to differentiate bacterial from viral infection in children with community-acquired pneumonia. BMC Pulmonary Medicine. 2016;16(1):103.

23. Romejko K, Markowska M, Niemczyk S. The Review of Current Knowledge on Neutrophil Gelatinase-Associated Lipocalin (NGAL). Int J Mol Sci. 2023;24(13).

24. Ashkenazi-Hoffnung L, Oved K, Navon R, Friedman T, Boico O, Paz M, et al. A host-protein signature is superior to other biomarkers for differentiating between bacterial and viral disease in patients with respiratory infection and fever without source: a prospective observational study. Eur J Clin Microbiol Infect Dis. 2018;37(7):1361–71.

25. Srugo I, Klein A, Stein M, Golan-Shany O, Kerem N, Chistyakov I, et al. Validation of a Novel Assay to Distinguish Bacterial and Viral Infections. Pediatrics. 2017;140(4).

26. Hainrichson M, Avni N, Eden E, Feigin P, Gelman A, Halabi S, et al. A point-of-need platform for rapid measurement of a host-protein score that differentiates bacterial from viral infection: Analytical evaluation. Clin Biochem. 2023;117:39–47.

27. Chokkalla AK, Tam E, Liang R, Cruz AT, Devaraj S. Validation of a multi-analyte immunoassay for distinguishing bacterial vs. viral infections in a pediatric cohort. Clin Chim Acta. 2023;546:117387.

28. Gillette MA, Mani DR, Uschnig C, Pellé KG, Madrid L, Acácio S, et al. Biomarkers to Distinguish Bacterial From Viral Pediatric Clinical Pneumonia in a Malaria-Endemic Setting. Clinical Infectious Diseases. 2021;73(11):e3939–e48.

29. Heinonen S, Rodriguez-Fernandez R, Diaz A, Oliva Rodriguez-Pastor S, Ramilo O, Mejias A. Infant Immune Response to Respiratory Viral Infections. Immunol Allergy Clin North Am. 2019;39(3):361–76.

30. Shaikh N, Campbell EA, Curry C, Mickles C, Cole EB, Liu H, et al. Accuracy of Screening Tests for the Diagnosis of Urinary Tract Infections in Young Children. Pediatrics. 2024.

31. Shaikh N, Shope TR, Hoberman A, Vigliotti A, Kurs-Lasky M, Martin JM. Association Between Uropathogen and Pyuria. Pediatrics. 2016;138(1).

32. Borensztajn DM, Zachariasse JM, Carrol ED, Nijman RG, von Both U, Emonts M, et al. Procalcitonin use in febrile children attending European emergency departments: a prospective multicenter study. BMC Pediatrics. 2025;25(1):157.

