## Supplemental Figures and Tables for "Proteomic Biomarkers Differentiate Bacterial Infections in Febrile Infants: A Multicentre Prospective Study"

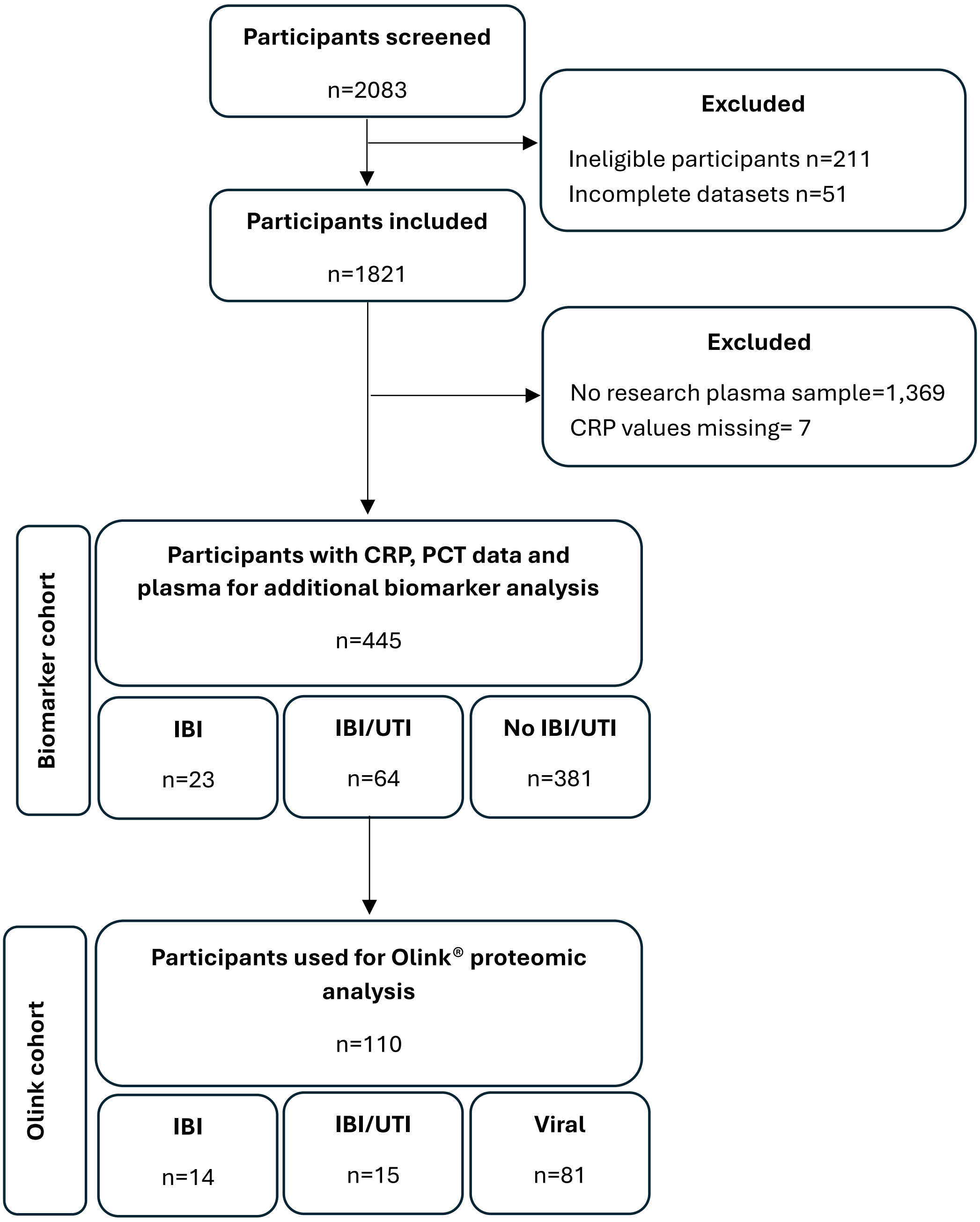


**Supplemental Figure 1.** Flow of participants through the study. *CRP, C-reactive protein; PCT, Procalcitonin; IBI, Invasive bacterial infection; UTI, Urinary tract infection.*

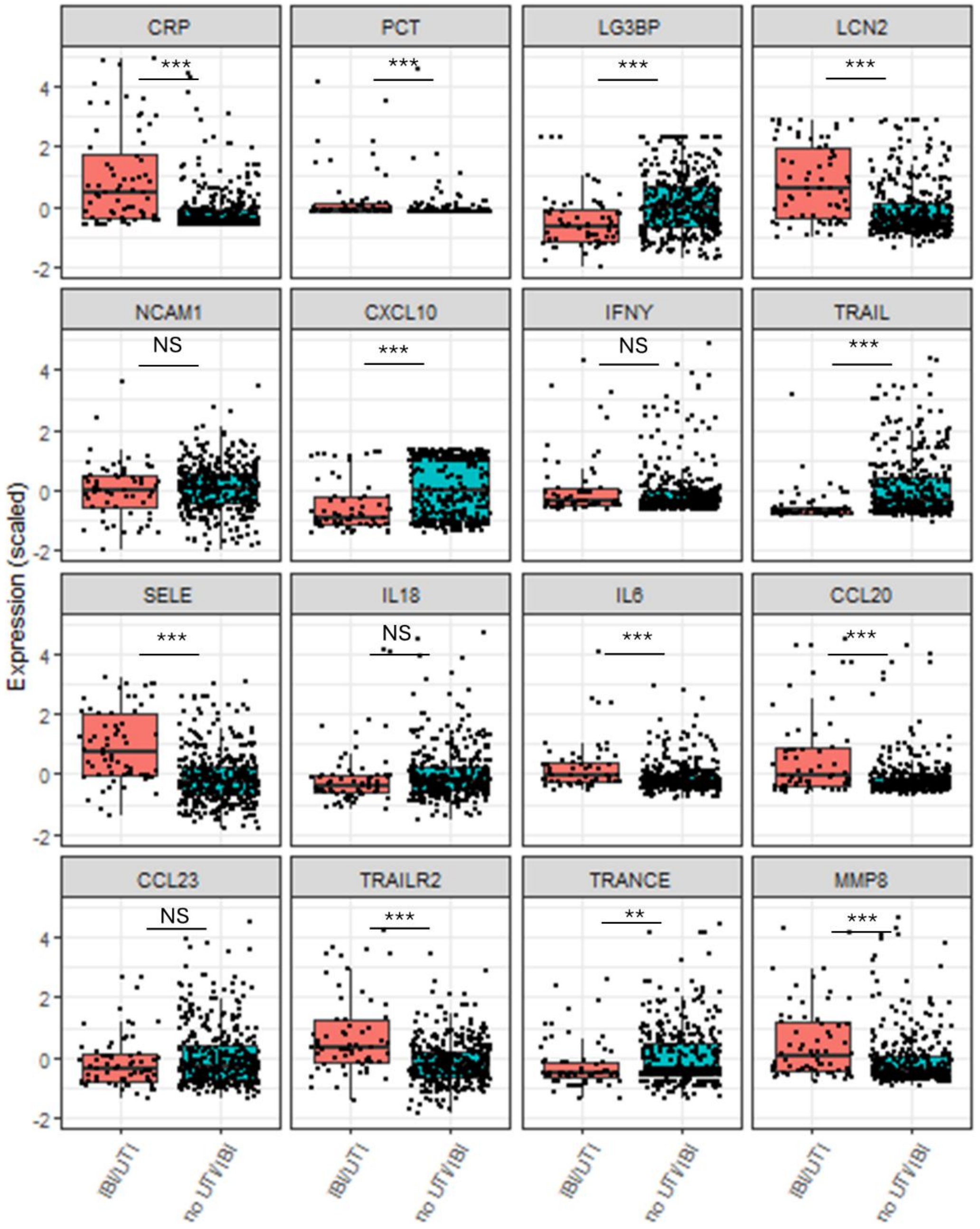


**Supplemental Figure 2. Box plots of Luminex and ELISA proteins in FIDO biomarker cohort with and without bacterial infection.** Box plots show scaled plasma protein expression values across samples from infants with and without UTI/IBI. P-values are displayed as; ns: p-value > 0.05; *: p-value <0.05; **: p-value < 0.01; ***: p-value ≤ 0.001.

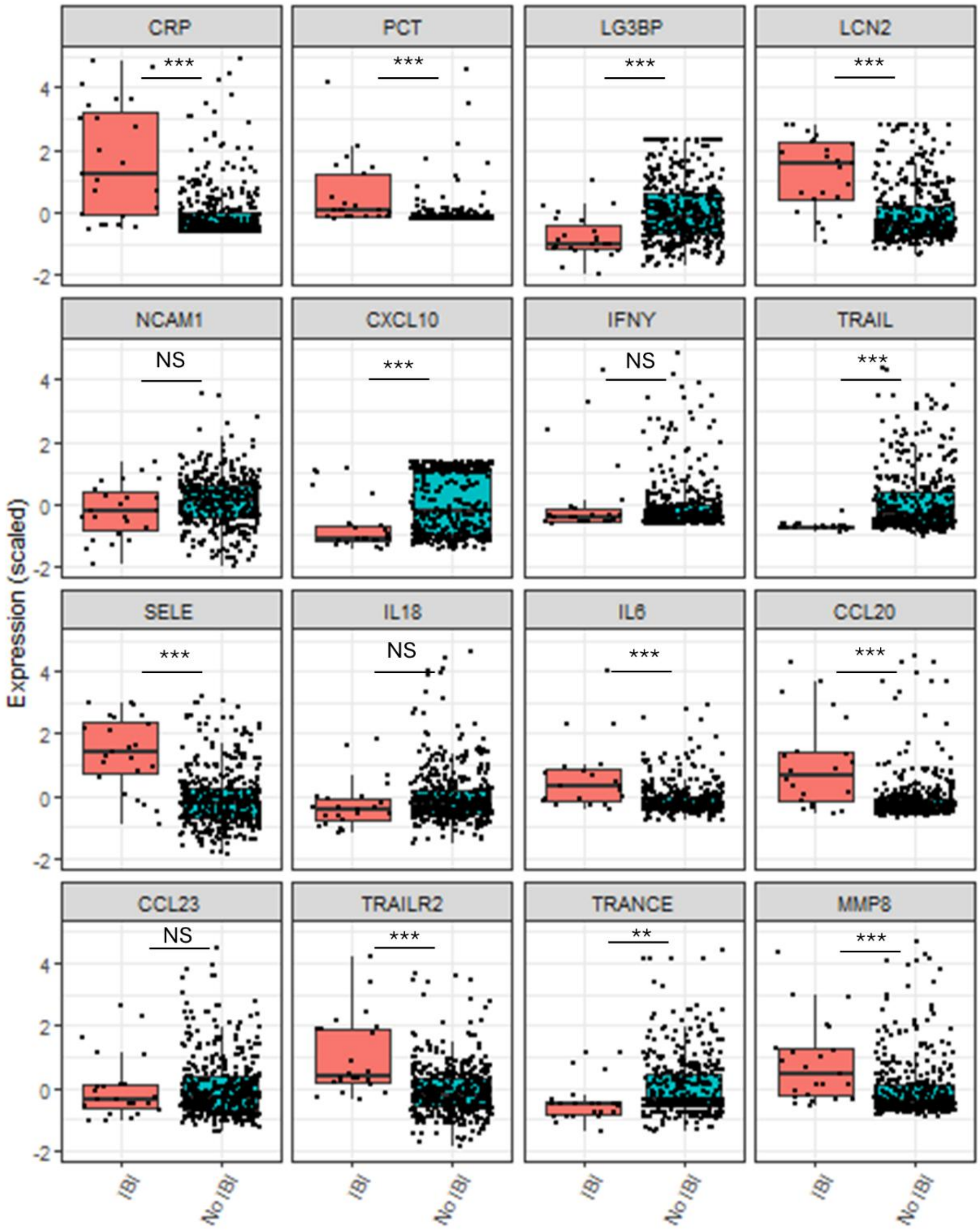


**Supplemental Figure 3. Box plots of Luminex and ELISA proteins in FIDO biomarker cohort with and without IBI.** Box plots show scaled plasma protein expression values across samples from infants with and without IBI. P-values are displayed as; ns: p-value > 0.05; *: p-value <0.05; **: p-value < 0.01; ***: p-value ≤ 0.001.

| | Biomarker cohort, n (%) | Total FIDO cohort, n (%) |
|---|---|---|
| **Positive blood cultures** | **22 (4.9)** | **62 (3.4)** |
| *Citrobacter freundii* | 0 (0) | 1 (1.6) |
| *Escherichia coli* | 11 (50) | 33 (53.2) |
| *Enterococcus spp·* | 0 (0) | 1 (1.6) |
| *Enterobacter spp.* | 0 (0) | 2 (3.2) |
| *Klebsiella pneumoniae* | 0 (0) | 1 (1.6) |
| *Neisseria meningitidis* | 2 (9.1) | 2 (3.2) |
| *Salmonella* | 0 (0) | 1 (1.6) |
| *Streptococcus agalactiae* | 3 (13.6) | 10 (16.1) |
| *Staphylococcus aureus* | 5 (22.7) | 9 (14.5) |
| *Streptococcus pyogenes* | 0 (0) | 1 (1.6) |
| *Streptococcus Gallolyticus* | 1 (4.5) | 1 (1.6) |
| **Positive CSF cultures** | **2 (0.4)** | **9 (0.5)** |
| *Escherichia coli* | 0 (0) | 1 (11.1) |
| *Enterococcus spp·* | 0 (0) | 1 (11.1) |
| *Enterobacter spp.* | 0 (0) | 1 (11.1) |
| *Neisseria meningitidis* | 1 (50) | 2 (22.2) |
| *Pseudomonas massiliensis* | 0 (0) | 1 (11.1) |
| *Streptococcus agalactiae* | 1 (50) | 3 (33.3) |
| **Positive urine cultures** | **45 (10.1)** | **147 (8.1)** |
| *Citrobacter freundii* | 0 (0) | 2 (1.4) |
| *Escherichia coli* | 35 (77.8) | 114 (77.6) |
| *Enterococcus spp·* | 7 (15.6) | 16 (10.9) |
| *Enterobacter cloacae* | 0 (0) | 1 (0.7) |
| *Klebsiella pneumoniae* | 1 (2.2) | 8 (5.4) |
| *Klebsiella oxytoca* | 1 (2.2) | 2 (1.4) |
| *Proteus mirablis* | 0 (0) | 2 (1.4) |
| *Streptococcus agalactiae* | 0 (0) | 1 (0.7) |
| *Staphylococcus aureus* | 1 (2.2) | 1 (0.7) |

**Supplemental Table 1**. Bacterial pathogen information for the full FIDO cohort and those available for additional biomarker testing. *CSF, Cerebrospinal fluid; spp., species.*

| Variable | Olink cohort |
|---|---|
| **n** | 110 |
| **Age (days), Median (IQR)** | 41 (26-59) |
| **Male, n(%)** | 74 (67.2) |
| **Positive blood cultures** | 13 |
| *Escherichia coli* | 8 |
| *Neisseria meningitidis* | 1 |
| *Streptococcus agalactiae* | 2 |
| *Staphylococcus aureus* | 1 |
| *Streptococcus Gallolyticus* | 1 |
| **Positive CSF cultures** | 2 |
| *Neisseria meningitidis* | 1 |
| *Streptococcus agalactiae* | 1 |
| **Positive urine cultures** | 18 |
| *Escherichia coli* | 13 |
| *Enterococcus spp·* | 3 |
| *Klebsiella oxytoca* | 1 |
| *Staphylococcus aureus* | 1 |

**Supplemental Table 2**. Olink cohort demographics. *CSF, Cerebrospinal fluid; IQR; Interquartile range.*

| Assay | OlinkID | UniProt | Panel | Fold Change | p.value | Adjusted pval |
|---|---|---|---|---|---|---|
| TNFSF11 | OID20592 | O14788 | Inflammation | -2.288 | <0.0001 | <0.0001 |
| C19orf12 | OID20804 | Q9NSK7 | Neurology | -1.225 | <0.0001 | <0.0001 |
| IL22RA1 | OID20449 | Q8N6P7 | Inflammation | -0.717 | <0.0001 | <0.0001 |
| TNFSF10 | OID20611 | P50591 | Inflammation | -1.196 | <0.0001 | <0.0001 |
| IFNL1 | OID20795 | Q8IU54 | Neurology | -2.967 | <0.0001 | <0.0001 |
| MMP8 | OID21019 | P22894 | Neurology | 1.424 | <0.0001 | <0.0001 |
| FCAR | OID20564 | P24071 | Inflammation | 1.325 | <0.0001 | 0.0001 |
| SUMF2 | OID21032 | Q8NBJ7 | Neurology | -0.608 | <0.0001 | 0.0001 |
| AGRN | OID20786 | O00468 | Inflammation | -0.749 | <0.0001 | 0.0002 |
| LAMP3 | OID20638 | Q9UQV4 | Inflammation | -1.010 | <0.0001 | 0.0002 |
| CCL20 | OID20671 | P78556 | Inflammation | 1.901 | <0.0001 | 0.0003 |
| NPPC | OID20568 | P23582 | Inflammation | -0.737 | <0.0001 | 0.0003 |
| FASLG | OID20665 | P48023 | Inflammation | -0.599 | <0.0001 | 0.0003 |
| GRN | OID21159 | P28799 | Neurology | -0.641 | <0.0001 | 0.0004 |
| SIGLEC1 | OID20690 | Q9BZZ2 | Inflammation | -0.740 | <0.0001 | 0.0004 |
| CALCA | OID20983 | P01258 | Neurology | 1.599 | <0.0001 | 0.0004 |
| TPSAB1 | OID20642 | Q15661 | Inflammation | -0.577 | <0.0001 | 0.0005 |
| SCARB2 | OID20943 | Q14108 | Neurology | -0.517 | <0.0001 | 0.0005 |
| SIGLEC10 | OID20537 | Q96LC7 | Inflammation | 0.892 | <0.0001 | 0.0006 |
| CLEC4D | OID20609 | Q8WXI8 | Inflammation | 0.966 | <0.0001 | 0.0006 |

| | | | | | | |
|---|---|---|---|---|---|---|
| TNFRSF10B | OID20981 | O14763 | Neurology | 0.764 | <0.0001 | 0.0008 |
| IL6 | OID20563 | P05231 | Inflammation | 2.513 | <0.0001 | 0.0008 |
| TGFA | OID20600 | P01135 | Inflammation | 1.051 | <0.0001 | 0.0008 |
| CCL23 | OID20693 | P55773 | Inflammation | 1.121 | <0.0001 | 0.0008 |
| LTA | OID20586 | P01374 | Inflammation | -0.539 | <0.0001 | 0.0008 |
| CXCL11 | OID21042 | O14625 | Neurology | -1.926 | <0.0001 | 0.0008 |
| KYNU | OID20669 | Q16719 | Inflammation | -0.486 | <0.0001 | 0.0010 |
| CLEC4C | OID20547 | Q8WTT0 | Inflammation | -0.998 | <0.0001 | 0.0010 |
| DSC2 | OID21079 | Q02487 | Neurology | 0.436 | <0.0001 | 0.0011 |
| CD274 | OID20966 | Q9NZQ7 | Neurology | -0.765 | <0.0001 | 0.0011 |
| CTSO | OID20660 | P43234 | Inflammation | -0.615 | <0.0001 | 0.0018 |
| LGALS9 | OID20781 | O00182 | Inflammation | -0.595 | <0.0001 | 0.0020 |
| CXCL10 | OID20697 | P02778 | Inflammation | -1.942 | 0.0001 | 0.0022 |
| SPOCK1 | OID20924 | Q08629 | Neurology | -0.393 | 0.0001 | 0.0022 |
| TNFRSF11B | OID20735 | O00300 | Inflammation | 0.846 | 0.0001 | 0.0027 |
| ERBB3 | OID20705 | P21860 | Inflammation | -0.302 | 0.0001 | 0.0029 |
| TMPRSS5 | OID20947 | Q9H3S3 | Neurology | -0.429 | 0.0002 | 0.0035 |
| LAMA4 | OID20769 | Q16363 | Inflammation | -0.584 | 0.0002 | 0.0038 |
| IL17A | OID20469 | Q16552 | Inflammation | 1.600 | 0.0002 | 0.0040 |
| CD34 | OID21025 | P28906 | Neurology | -0.288 | 0.0002 | 0.0043 |
| SPRY2 | OID20475 | O43597 | Inflammation | -0.680 | 0.0003 | 0.0053 |
| ADAM23 | OID20651 | O75077 | Inflammation | -0.336 | 0.0003 | 0.0057 |
| IFNLR1 | OID20506 | Q8IU57 | Inflammation | -0.838 | 0.0003 | 0.0057 |
| TLR3 | OID20612 | O15455 | Inflammation | -0.808 | 0.0004 | 0.0060 |
| SFRP1 | OID20984 | Q8N474 | Neurology | -0.595 | 0.0004 | 0.0070 |
| ISLR2 | OID20889 | Q6UXK2 | Neurology | -0.421 | 0.0005 | 0.0070 |
| CD244 | OID20628 | Q9BZW8 | Inflammation | -0.283 | 0.0005 | 0.0071 |
| MDGA1 | OID20951 | Q8NFP4 | Neurology | -0.711 | 0.0005 | 0.0071 |
| CRLF1 | OID20699 | O75462 | Inflammation | -0.322 | 0.0005 | 0.0074 |
| OBP2B | OID20980 | Q9NPH6 | Neurology | -0.661 | 0.0005 | 0.0074 |
| GPC5 | OID20944 | P78333 | Neurology | -0.421 | 0.0006 | 0.0078 |
| GZMA | OID20663 | P12544 | Inflammation | -0.426 | 0.0006 | 0.0078 |
| CXCL17 | OID20622 | Q6UXB2 | Inflammation | -0.560 | 0.0006 | 0.0078 |
| IL32 | OID20605 | P24001 | Inflammation | -0.486 | 0.0006 | 0.0078 |
| LGMN | OID20773 | Q99538 | Inflammation | -0.422 | 0.0006 | 0.0078 |
| BAG3 | OID20972 | O95817 | Neurology | -0.405 | 0.0006 | 0.0079 |
| RGMA | OID21065 | Q96B86 | Neurology | -0.235 | 0.0006 | 0.0079 |
| NME3 | OID20789 | Q13232 | Inflammation | -0.298 | 0.0006 | 0.0079 |
| CHRDL1 | OID20771 | Q9BU40 | Inflammation | -0.422 | 0.0007 | 0.0079 |
| MVK | OID20459 | Q03426 | Inflammation | -0.696 | 0.0007 | 0.0079 |
| FGF5 | OID20490 | P12034 | Inflammation | -0.348 | 0.0007 | 0.0079 |
| CSF3 | OID20491 | P09919 | Inflammation | 1.426 | 0.0007 | 0.0080 |
| BTN2A1 | OID20713 | Q7KYR7 | Inflammation | -0.292 | 0.0007 | 0.0084 |
| CCL3 | OID20610 | P10147 | Inflammation | 1.144 | 0.0008 | 0.0084 |
| DNER | OID20712 | Q8NFT8 | Inflammation | -0.238 | 0.0008 | 0.0092 |
| STC2 | OID21157 | O76061 | Neurology | -0.313 | 0.0010 | 0.0103 |

| | | | | | | |
|---|---|---|---|---|---|---|
| LRRN1 | OID20438 | Q6UXK5 | Inflammation | -0.336 | 0.0010 | 0.0103 |
| WARS1 | OID21084 | P23381 | Neurology | -0.707 | 0.0010 | 0.0105 |
| EPO | OID20522 | P01588 | Inflammation | 0.998 | 0.0010 | 0.0106 |
| IL7 | OID20535 | P13232 | Inflammation | -0.595 | 0.0011 | 0.0111 |
| CCL4 | OID20695 | P13236 | Inflammation | 1.287 | 0.0011 | 0.0115 |
| TNFSF14 | OID20953 | O43557 | Neurology | 0.534 | 0.0012 | 0.0115 |
| CDH15 | OID21005 | P55291 | Neurology | -0.459 | 0.0012 | 0.0115 |
| RGMB | OID21045 | Q6NW40 | Neurology | -0.273 | 0.0012 | 0.0117 |
| PROK1 | OID20543 | P58294 | Inflammation | 0.489 | 0.0012 | 0.0117 |
| MEGF10 | OID20746 | Q96KG7 | Inflammation | -0.324 | 0.0012 | 0.0117 |
| TNFRSF1A | OID21155 | P19438 | Neurology | 0.604 | 0.0013 | 0.0117 |
| ROBO2 | OID21007 | Q9HCK4 | Neurology | -0.344 | 0.0013 | 0.0117 |
| FCRL6 | OID20529 | Q6DN72 | Inflammation | -0.647 | 0.0013 | 0.0117 |
| CD109 | OID21108 | Q6YHK3 | Neurology | -0.285 | 0.0014 | 0.0121 |
| VASH1 | OID20516 | Q7L8A9 | Inflammation | -0.369 | 0.0014 | 0.0126 |
| HLA-E | OID20532 | P13747 | Inflammation | -0.424 | 0.0015 | 0.0126 |
| BSG | OID20626 | P35613 | Inflammation | -0.212 | 0.0015 | 0.0126 |
| CST7 | OID20704 | O76096 | Inflammation | 0.754 | 0.0015 | 0.0126 |
| EPCAM | OID20743 | P16422 | Inflammation | -0.407 | 0.0015 | 0.0126 |
| DECR1 | OID20579 | Q16698 | Inflammation | -0.828 | 0.0015 | 0.0126 |
| IL11 | OID20455 | P20809 | Inflammation | 0.853 | 0.0015 | 0.0126 |
| TST | OID20868 | Q16762 | Neurology | -0.314 | 0.0015 | 0.0126 |
| CA6 | OID21096 | P23280 | Neurology | -0.771 | 0.0016 | 0.0127 |
| NGF | OID20920 | P01138 | Neurology | -0.056 | 0.0017 | 0.0133 |
| TREM2 | OID20731 | Q9NZC2 | Inflammation | -0.404 | 0.0018 | 0.0140 |
| SCG3 | OID20728 | Q8WXD2 | Inflammation | -0.412 | 0.0018 | 0.0142 |
| GALNT3 | OID20471 | Q14435 | Inflammation | -0.322 | 0.0019 | 0.0145 |
| AGR2 | OID20896 | O95994 | Neurology | -0.712 | 0.0019 | 0.0145 |
| MERTK | OID20657 | Q12866 | Inflammation | -0.278 | 0.0021 | 0.0160 |
| VSTM1 | OID20851 | Q6UX27 | Neurology | 0.632 | 0.0021 | 0.0160 |
| AGER | OID20756 | Q15109 | Inflammation | -0.371 | 0.0022 | 0.0161 |
| CRHBP | OID20747 | P24387 | Inflammation | 0.455 | 0.0022 | 0.0162 |
| NTRK3 | OID21057 | Q16288 | Neurology | -0.233 | 0.0023 | 0.0167 |
| IL24 | OID20440 | Q13007 | Inflammation | 0.912 | 0.0024 | 0.0168 |
| PTX3 | OID20570 | P26022 | Inflammation | 0.430 | 0.0024 | 0.0168 |
| CDON | OID20754 | Q4KMG0 | Inflammation | -0.409 | 0.0024 | 0.0170 |
| RHOC | OID20950 | P08134 | Neurology | -0.580 | 0.0025 | 0.0174 |
| OSM | OID20574 | P13725 | Inflammation | 0.952 | 0.0025 | 0.0174 |
| PRTFDC1 | OID20867 | Q9NRG1 | Neurology | -0.769 | 0.0025 | 0.0174 |
| CNTN3 | OID21092 | Q9P232 | Neurology | -0.353 | 0.0026 | 0.0179 |
| IL10 | OID20431 | P22301 | Inflammation | 1.868 | 0.0027 | 0.0180 |
| FSTL3 | OID20782 | O95633 | Inflammation | 0.635 | 0.0027 | 0.0181 |
| CCL25 | OID20674 | O15444 | Inflammation | -0.429 | 0.0028 | 0.0184 |
| SUSD2 | OID21098 | Q9UGT4 | Neurology | -0.383 | 0.0028 | 0.0184 |
| LIF | OID20824 | P15018 | Neurology | 1.524 | 0.0029 | 0.0187 |
| DKK4 | OID20934 | Q9UBT3 | Neurology | -0.523 | 0.0029 | 0.0187 |

| | | | | | | |
|---|---|---|---|---|---|---|
| SEMA4D | OID21020 | Q92854 | Neurology | -0.375 | 0.0029 | 0.0187 |
| PRSS8 | OID20763 | Q16651 | Inflammation | -0.284 | 0.0030 | 0.0187 |
| GZMB | OID20604 | P10144 | Inflammation | -0.825 | 0.0030 | 0.0187 |
| IL33 | OID20428 | O95760 | Inflammation | -0.603 | 0.0032 | 0.0197 |
| LAP3 | OID20436 | P28838 | Inflammation | -0.530 | 0.0032 | 0.0197 |
| KLRB1 | OID20629 | Q12918 | Inflammation | -0.376 | 0.0033 | 0.0199 |
| NCAN | OID21055 | O14594 | Neurology | -0.370 | 0.0034 | 0.0204 |
| CD58 | OID20716 | P19256 | Inflammation | -0.141 | 0.0034 | 0.0204 |
| TNFRSF1B | OID21145 | P20333 | Neurology | 0.499 | 0.0036 | 0.0212 |
| ITGA11 | OID20581 | Q9UKX5 | Inflammation | -0.605 | 0.0036 | 0.0212 |
| CPA2 | OID21060 | P48052 | Neurology | -0.585 | 0.0036 | 0.0212 |
| ECE1 | OID20891 | P42892 | Neurology | -0.169 | 0.0038 | 0.0219 |
| CTSS | OID21056 | P25774 | Neurology | -0.346 | 0.0039 | 0.0223 |
| FCRL2 | OID20639 | Q96LA5 | Inflammation | -0.589 | 0.0039 | 0.0223 |
| HNMT | OID20975 | P50135 | Neurology | -0.548 | 0.0040 | 0.0225 |
| TNF | OID20473 | P01375 | Inflammation | 0.959 | 0.0040 | 0.0226 |
| FKBP7 | OID20846 | Q9Y680 | Neurology | -0.289 | 0.0041 | 0.0230 |
| B4GALT1 | OID20780 | P15291 | Inflammation | 0.282 | 0.0043 | 0.0240 |
| SLC39A5 | OID20539 | Q6ZMH5 | Inflammation | -0.557 | 0.0044 | 0.0243 |
| SOD2 | OID21114 | P04179 | Neurology | -0.338 | 0.0045 | 0.0247 |
| IL12RB1 | OID20486 | P42701 | Inflammation | -0.396 | 0.0046 | 0.0251 |
| GOLM2 | OID21044 | Q6P4E1 | Neurology | -0.186 | 0.0049 | 0.0261 |
| BMP4 | OID20994 | P12644 | Neurology | -0.607 | 0.0049 | 0.0261 |
| CD200R1 | OID20595 | Q8TD46 | Inflammation | -0.303 | 0.0051 | 0.0268 |
| B4GAT1 | OID21127 | O43505 | Neurology | -0.289 | 0.0051 | 0.0269 |
| ALDH1A1 | OID21128 | P00352 | Neurology | -0.542 | 0.0054 | 0.0280 |
| SMOC2 | OID20730 | Q9H3U7 | Inflammation | -0.246 | 0.0055 | 0.0281 |
| IL2 | OID20419 | P60568 | Inflammation | 0.796 | 0.0057 | 0.0293 |
| NELL2 | OID20706 | Q99435 | Inflammation | -0.234 | 0.0058 | 0.0295 |
| SCARF2 | OID21048 | Q96GP6 | Neurology | -0.312 | 0.0063 | 0.0319 |
| FABP9 | OID20501 | Q0Z7S8 | Inflammation | -0.570 | 0.0064 | 0.0323 |
| ITGA6 | OID20528 | P23229 | Inflammation | -0.512 | 0.0065 | 0.0323 |
| BCAN | OID20998 | Q96GW7 | Neurology | -0.443 | 0.0067 | 0.0333 |
| PRKAB1 | OID20444 | Q9Y478 | Inflammation | -0.341 | 0.0069 | 0.0340 |
| CD276 | OID20680 | Q5ZPR3 | Inflammation | -0.299 | 0.0070 | 0.0342 |
| ISM1 | OID20538 | B1AKI9 | Inflammation | -0.326 | 0.0072 | 0.0349 |
| ADAM22 | OID21001 | Q9P0K1 | Neurology | -0.283 | 0.0072 | 0.0349 |
| TRIM21 | OID20533 | P19474 | Inflammation | -0.533 | 0.0075 | 0.0361 |
| HPCAL1 | OID20615 | P37235 | Inflammation | -0.388 | 0.0076 | 0.0363 |
| VSIG4 | OID21144 | Q9Y279 | Neurology | 0.518 | 0.0081 | 0.0380 |
| WAS | OID20479 | P42768 | Inflammation | -0.354 | 0.0084 | 0.0388 |
| KLB | OID20888 | Q86Z14 | Neurology | -0.715 | 0.0084 | 0.0388 |
| HGF | OID20656 | P14210 | Inflammation | 0.639 | 0.0084 | 0.0388 |
| CNTN5 | OID21006 | O94779 | Neurology | -0.325 | 0.0084 | 0.0388 |
| LAT | OID20640 | O43561 | Inflammation | -0.512 | 0.0087 | 0.0399 |
| IL2RB | OID20450 | P14784 | Inflammation | -0.280 | 0.0091 | 0.0413 |

| | | | | | | |
|---|---|---|---|---|---|---|
| PCDH1 | OID20614 | Q08174 | Inflammation | -0.124 | 0.0095 | 0.0427 |
| NTF3 | OID20591 | P20783 | Inflammation | -0.366 | 0.0095 | 0.0427 |
| ENAH | OID20497 | Q8N8S7 | Inflammation | -0.373 | 0.0097 | 0.0434 |
| CD48 | OID20692 | P09326 | Inflammation | -0.286 | 0.0102 | 0.0454 |
| NCK2 | OID20683 | O43639 | Inflammation | -0.464 | 0.0105 | 0.0463 |
| TNFSF12 | OID20624 | O43508 | Inflammation | -0.393 | 0.0110 | 0.0482 |
| BST2 | OID21029 | Q10589 | Neurology | -0.581 | 0.0111 | 0.0485 |
| MPO | OID21100 | P05164 | Neurology | 0.596 | 0.0112 | 0.0486 |
| MESD | OID21099 | Q14696 | Neurology | -0.504 | 0.0113 | 0.0486 |
| FUT8 | OID20992 | Q9BYC5 | Neurology | -0.479 | 0.0116 | 0.0492 |
| CD79B | OID20635 | P40259 | Inflammation | -0.536 | 0.0116 | 0.0492 |
| GFRA3 | OID20933 | O60609 | Neurology | -0.277 | 0.0117 | 0.0492 |
| IL1B | OID20427 | P01584 | Inflammation | 1.199 | 0.0118 | 0.0495 |
| PON3 | OID20777 | Q15166 | Inflammation | -0.404 | 0.0119 | 0.0495 |
| NRP2 | OID21081 | O60462 | Neurology | -0.217 | 0.0120 | 0.0499 |
| LRPAP1 | OID21059 | P30533 | Neurology | -0.441 | 0.0121 | 0.0499 |

**Supplemental Table 3**. Differentially expressed plasma proteins between infants in Olink cohort, with bacterial or viral infection (FDR<0.05).

| **Biomarker** | | **Uniprot** | **Platform measured** | **Reason for inclusion** | **Infection type raised** |
|---|---|---|---|---|---|
| **TRAIL** | TNF-related apoptosis-inducing ligand | P50591 | Luminex | Olink and Literature | Viral |
| **SELE** | E-selectin | P16581 | Luminex | Literature | Bacterial |
| **LCN2** | Lipocalin 2 | P80188 | ELISA | Literature | Bacterial |
| **IL6** | Interleukin-6 | P05231 | Luminex | Olink | Bacterial |
| **TRAILR2** | TRAIL receptor 2 | O14763 | Luminex | Olink | Bacterial |
| **CCL20** | C-C motif chemokine 20 | P78556 | Luminex | Olink | Bacterial |
| **MMP8** | matrix metalloproteinase-8 | P22894 | Luminex | Olink | Bacterial |
| **LG3BP** | Galectin-3-binding protein | Q08380 | ELISA | Literature | Viral |
| **CXCL10** | C-X-C motif chemokine 10 | P02778 | Luminex | Olink and Literature | Viral |
| **TRANCE** | Tumor necrosis factor ligand superfamily member 11 | O14788 | Luminex | Olink | Viral |
| **NCAM1** | Neural cell adhesion molecule 1 | P13591 | Luminex | Literature | Viral |
| **IL18** | Interleukin-18 | Q14116 | Luminex | Literature | Viral |
| **IL17A** | Interleukin-17A | Q16552 | Luminex | Olink | Bacterial |
| **IFNG** | Interferon gamma | P01579 | Luminex | Literature | Viral |
| **CCL23** | C-C motif chemokine 23 | P55773 | Luminex | Olink | Bacterial |

**Supplemental Table 4**. Proteins selected for measurement in full biomarker cohort.

| Biomarker | IBI, AUC (95% CI) | IBI/UTI, AUC (95% CI) |
|---|---|---|
| CRP | 0.82 (0.73, 0.91) | 0.75 (0.67, 0.82) |
| PCT | 0.89 (0.82, 0.97) | 0.76 (0.7, 0.83) |
| Oved et al | 0.89 (0.83, 0.95) | 0.78 (0.72, 0.84) |
| TRAIL | 0.86 (0.8, 0.91) | 0.75 (0.69, 0.81) |
| SELE | 0.83 (0.73, 0.92) | 0.77 (0.71, 0.84) |
| LCN2 | 0.81 (0.72, 0.91) | 0.73 (0.66, 0.8) |
| IL6 | 0.81 (0.72, 0.9) | 0.71 (0.65, 0.78) |
| TRAILR2 | 0.8 (0.72, 0.88) | 0.73 (0.66, 0.8) |
| CCL20 | 0.78 (0.66, 0.89) | 0.69 (0.61, 0.77) |
| MMP8 | 0.76 (0.67, 0.85) | 0.68 (0.61, 0.75) |
| LG3BP | 0.76 (0.66, 0.86) | 0.69 (0.62, 0.76) |
| CXCL10 | 0.73 (0.63, 0.83) | 0.68 (0.61, 0.75) |
| Jackson et al Simple | 0.68 (0.56, 0.8) | 0.68 (0.6, 0.75) |
| TRANCE | 0.66 (0.54, 0.78) | 0.6 (0.53, 0.68) |
| Jackson et al Weighted | 0.6 (0.49, 0.72) | 0.54 (0.46, 0.61) |
| NCAM1 | 0.58 (0.45, 0.71) | 0.54 (0.46, 0.62) |
| IL18 | 0.58 (0.44, 0.72) | 0.56 (0.48, 0.64) |
| IFNY | 0.53 (0.41, 0.66) | 0.53 (0.45, 0.61) |
| CCL23 | 0.5 (0.38, 0.63) | 0.54 (0.46, 0.61) |

**Supplemental Table 5.** AUC values for each biomarker to distinguish IBI from non-IBI infections, or IBI/UTI (all bacterial infections) from non-bacterial infections.

| Variable | Level | Estimate | Odds Ratio | StdErr | P.Value |
|---|---|---|---|---|---|
| | | | **UTI/IBI** | | |
| Intercept | | -1.7324 | 0.177 (0.090, 0.339) | 0.3362 | <1e-04 *** |
| CRP | | 0.0172 | 1.017 (1.010, 1.025) | 0.0038 | <1e-04 *** |
| TRAIL | | -0.0037 | 0.996 (0.993, 0.999) | 0.0014 | 0.011 * |
| | | | **IBI** | | |
| Intercept | | -2.3480 | 0.096 (0.025, 0.340) | 0.6582 | 0.0004 *** |
| CRP | | 0.0160 | 1.016 (1.007, 1.025) | 0.0045 | 0.0004 *** |
| TRAIL | | -0.0116 | 0.988 (0.975, 0.997) | 0.0058 | 0.0445 * |

**Supplemental Table 6.** Multivariable binomial logistic model output for predicting IBI/UTI and IBI with CRP and TRAIL as variables.
